# The Impact of the Covid-19 Pandemic on Uptake of Influenza Vaccine: A UK-Wide Observational Study

**DOI:** 10.1101/2020.10.01.20205385

**Authors:** Patrik Bachtiger, Alex Adamson, Ji-Jian Chow, Rupa Sisodia, Jennifer K Quint, Nicholas S Peters

**Author notes:** The Corresponding Author has the right to grant on behalf of all authors and does grant on behalf of all authors, a worldwide licence to the Publishers and its licensees in perpetuity, in all forms, formats and media (whether known now or created in the future), to i) publish, reproduce, distribute, display and store the Contribution, ii) translate the Contribution into other languages, create adaptations, reprints, include within collections and create summaries, extracts and/or, abstracts of the Contribution, iii) create any other derivative work(s) based on the Contribution, iv) to exploit all subsidiary rights in the Contribution, v) the inclusion of electronic links from the Contribution to third party material where-ever it may be located; and, vi) licence any third party to do any or all of the above.

## Abstract

**Objectives:** The objective of this study was to measure the impact of the Covid-19 pandemic on acceptance of flu vaccination in the 2020-21 season, including for those newly eligible for the UK National Health Service (NHS) free vaccination programme, extended this year to include an estimated 32.4 million (48.8%) of the UK population. Knowing intended uptake is essential to inform supply and steer public health messaging to maximise vaccination given the combined threats of both flu and Covid-19 — the unknown impact of which on both attitudes and the need for mass uptake yet again create the threat of ill-informed planning resulting in failure to meet necessary public health demand.

**Methods:** An online questionnaire posing question items on influenza vaccination was administered to registrants of the Care Information Exchange (CIE), the NHS’s largest patient electronic personal health record. This was part of a longitudinal study initiated during the Covid-19 pandemic lockdown. This analysis was limited to those who, in line with established NHS criteria, were previously or newly eligible but had not routinely received seasonal influenza vaccination in the past. Groups were stratified by response (yes/no) to intending to receive flu vaccination in 2020-21: Group 1.) Previously eligible now responding ‘yes’, 2.) Previously eligible still responding ‘no’, 3.) Newly eligible responding ‘yes’, and 4.) Newly eligible responding ‘no’. Within these groups, response by health worker status and each group’s inclination to vaccinate school age children was also measured. Summary statistics were reported alongside univariate and multivariable regression. Lastly, a network analysis visualised the frequency and co-occurrence of reasons qualifying response for or against influenza vaccination in 2020/21.

**Findings:** Among 6,641 respondents, 4,040 (61.1%) had previously routinely received the flu vaccination. 1,624 (24.5%) had been either previously eligible but not vaccinated (945, 58.2%) or newly eligible (679, 41.8%). Among the previously eligible participants who had not routinely received influenza vaccination 536 (56.7%) responded they would in 2020-21, increasing the vaccination rate in the entire previously eligible cohort from 79.6% to 91.2%, and 466 (68.6%) in the newly eligible.

Multivariable logistic regression resulted in few substantial changes to effect estimates, with the exception of age, for which all estimates showed a stronger association with intention to receive the flu vaccine. In those who became newly eligible to receive the flu vaccine, there was an association between intention to receive the flu vaccine and increased age (OR = 1.07, 95% CI 1.03 to 1.12), IMD quintile, and considering oneself at high risk from Covid-19 (OR = 1.80, 95% CI 1.22 to 2.70).

Network analysis showed the most frequent themes for previously eligible unvaccinated and newly eligible participants accepting vaccination in 2020/21 were: ‘precaution for myself’ (41.2% and 46.1%) and ‘Covid-19’ (27.4% and 27.1%), where the former was qualified by the latter in 36% and 29.1% of responses. Among the previously and newly eligible not intending to receive vaccination in 2020/21, misinformed themes of ‘makes me unwell’, ‘gives me flu’ and ‘vaccine doesn’t work’ were present across 37.4% and 21.9% of responses, respectively.

Among participants with school age children, of those previously eligible who now intend to be vaccinated themselves, 82.5% also intend to vaccinate their children in 2020/21 compared to 25.8% of those who would not accept the influenza vaccine for themselves. Among the newly eligible respondents this was 82.1% and 43.5%, respectively. 49.9% of the previously unvaccinated healthcare workers would continue to decline the vaccine in 2020/21.

**Interpretation:** In this UK-wide observational study, Covid-19 has increased acceptance of flu vaccination in 2020/21 from 79.6% to 91.2% in those previously eligible, and for the 69% of those newly eligible. This high anticipated vaccination rate (to 26 million (80%) of the UK population) requires appropriate planning, but can be further increased with effective messaging campaigns to address negative misconceptions about flu vaccination, which may also help in preparation for future Covid-19 vaccination. It remains of concern that 50% of healthcare professionals who refused it previously still do not intend to have the flu vaccine.

## INTRODUCTION

Seasonal influenza (flu) puts the UK National Health Service (NHS) under considerable pressure each winter. The challenge of maximal bed occupancy at the peak of winter pressures is routinely compounded by up to 18,000 additional daily emergency admissions,^1^ with >4,000 hospital beds occupied by patients with flu during the peak of the severe outbreak in 2017/18,^2^ and corresponding pressures also impacting primary care.^3^

The coronavirus disease 2019 (Covid-19) pandemic, caused by the severe acute respiratory syndrome coronavirus 2 (SARS-CoV-2) virus, has thus far led to nearly 42,000 deaths in the UK alone. With increasing regional outbreaks,^4^ substantial concern has been raised about preparedness for a nationwide escalation of cases throughout winter pressures in 2020/21.^5–7^ For this reason, the NHS has extended its free seasonal flu vaccination programme to all people aged over 50 (previously 65), and to an expanded school-age range to include 11-12 year olds (previously 2-10 year olds).^8^ This expanded programme now makes an estimated 32.4 million (48.8%) of the UK population eligible,^9^ intended to minimise the burden of flu cases in a health service preparing for ongoing waves of Covid-19.

Early vaccination against influenza viruses is a cost-effective solution to prevent contagion and reduce the number of flu-related deaths,^10^ but in England in 2019 uptake among those eligible was only 70.6%,^11^ below the critical 75% target for effectiveness recommended by the World Health Organisation (WHO).^12^ On a background of declining numbers over the last decade, uptake this influenza season, 2020/21, is not only unknown but completely unpredictable. The threat of Covid-19 and the associated publicity educating the public about viruses and vaccine development, coupled with recent evidence that co-infection with influenza and SARS-CoV-2 doubles mortality compared with infection with Covid-19 alone,^13^ and that the flu vaccination may confer significant protection against Covid-19,^34^ are likely to affect attitudes and the public health imperative of mass uptake. With substantial concerns that higher earlier uptake of flu vaccination in 2020/21 will rapidly deplete stocks (already reported^14^) there is yet again a threat of a lack of informed planning resulting in failure to meet the demands of a public health initiative.

The objective of this study was to measure the impact of the Covid-19 pandemic on acceptance of flu vaccination in the 2020-21 season, including for those newly eligible. Knowing this is essential to inform supply and steer public health messaging to maximise uptake and help the NHS contend with a potential double winter pandemic of flu and Covid-19.

## METHODS

### Study Participants

Participants in this study were registrants of the Care Information Exchange (CIE) of Imperial College Healthcare NHS Foundation Trust (UK). The CIE is the UK’s largest patient-facing electronic health record, since 2016 accessible by email registration for any patient who has had an encounter (e.g. appointment, blood test, procedure) at the Trust. On the 5th of June 2020 the CIE held 57,056 registrants. Established registrant postcode data highlights that this is a UK-wide population (supplementary figure 1 showing map). The CIE documents age and sex for all registrants.

Participants in this study were CIE registrants receiving weekly questionnaires by email notification, starting 9th April 2020 (week 1), as a direct care tool for self-monitoring physical, mental and social wellbeing during the Covid-19 pandemic.

### Questionnaire Design & Timing

A questionnaire including items on the government’s expanded flu vaccination programme was sent to participants on 31st July 2020 (week 16). Applying recommendations for questionnaire design,^15,16^ question items were developed by a collaboration of experts in qualitative research at Imperial College London, encompassing public health, respiratory epidemiology and digital health, and were also informed by previous studies.^17,18^ Question items were externally peer-reviewed and tested on lay persons (n = 5) before being included in the final questionnaire. The focus was on previous uptake of flu vaccination, being for or against vaccination in 2020/21 and reasons why (unrestricted free text responses), health worker status and presence of school-age children in the household. The week 16 question items are included in supplementary materials table 1.

**Table 1:**
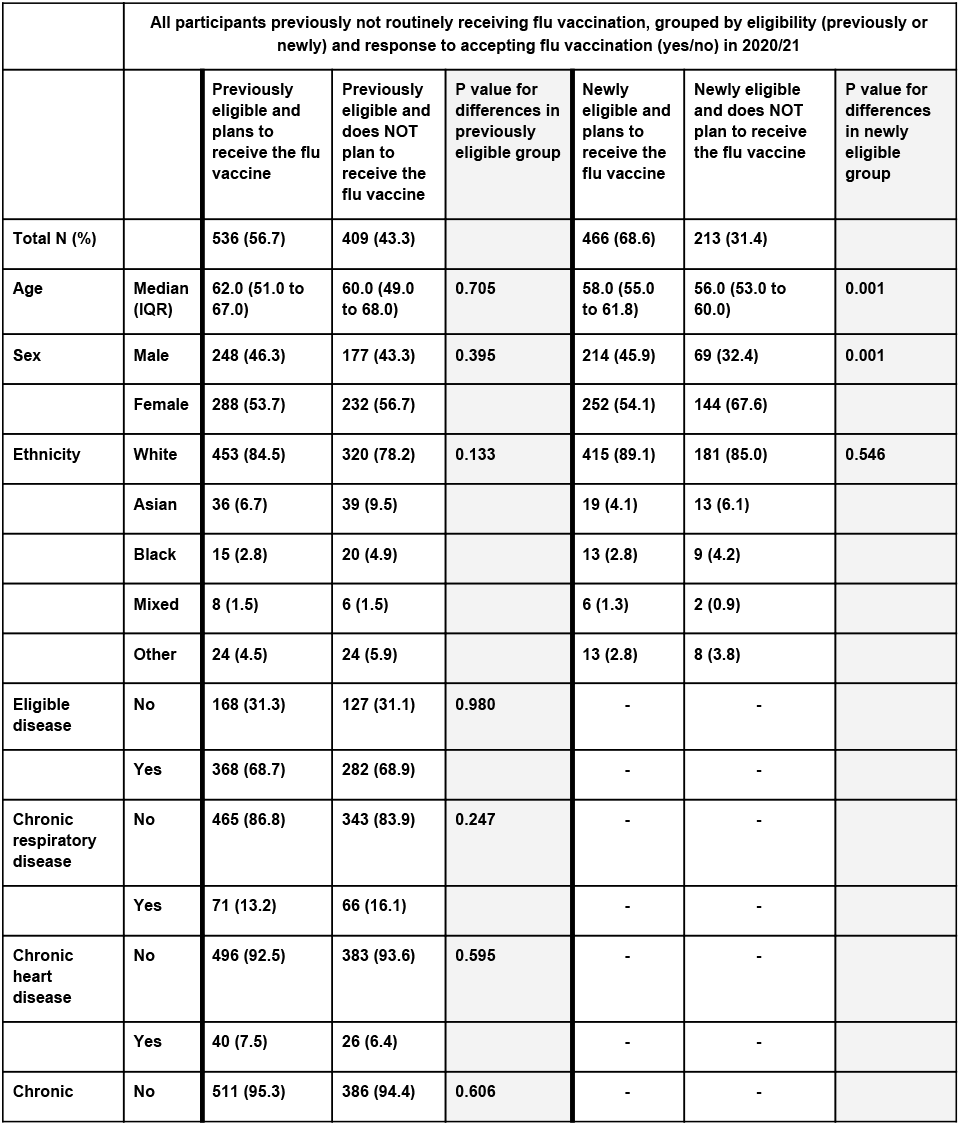
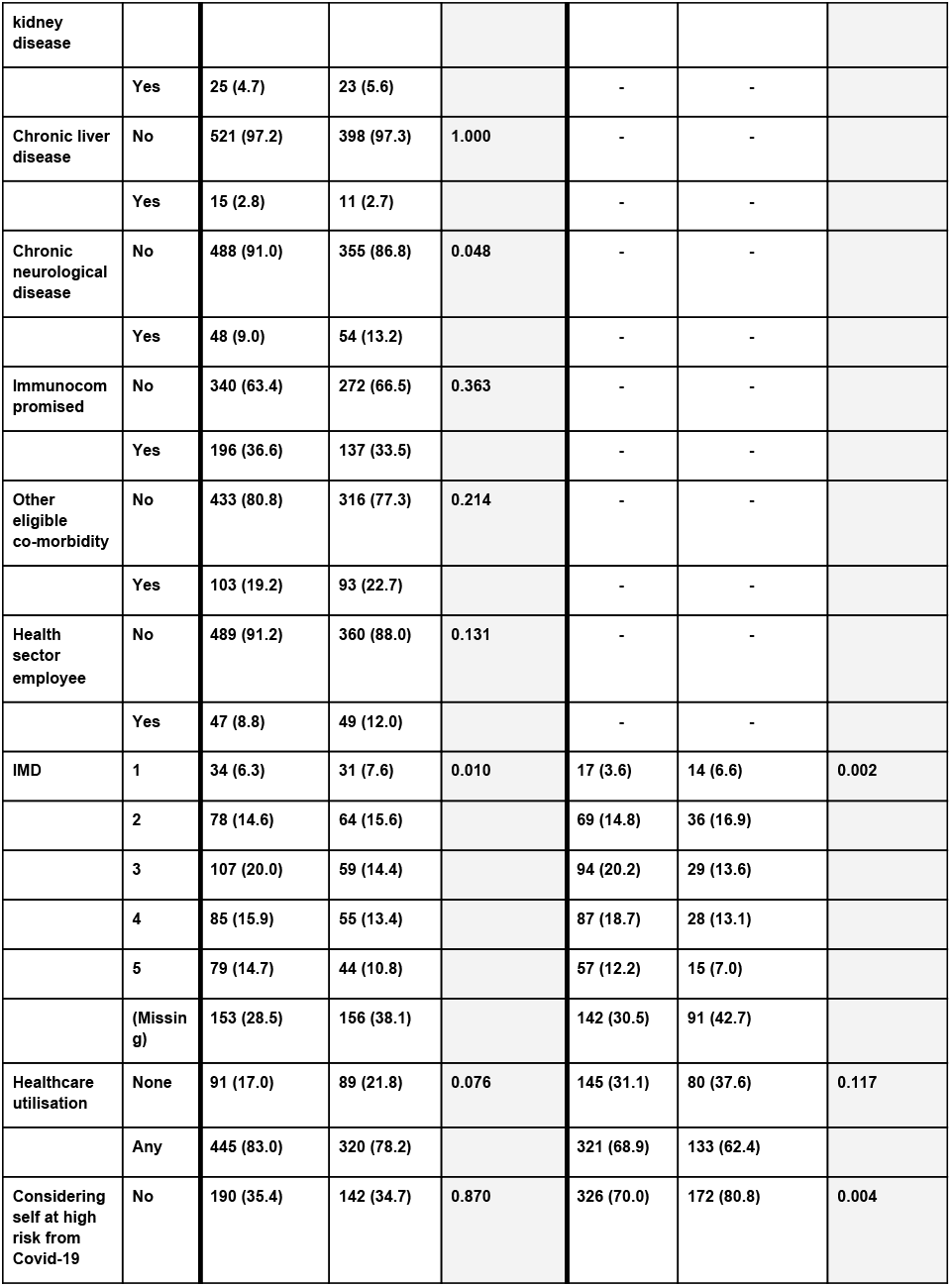
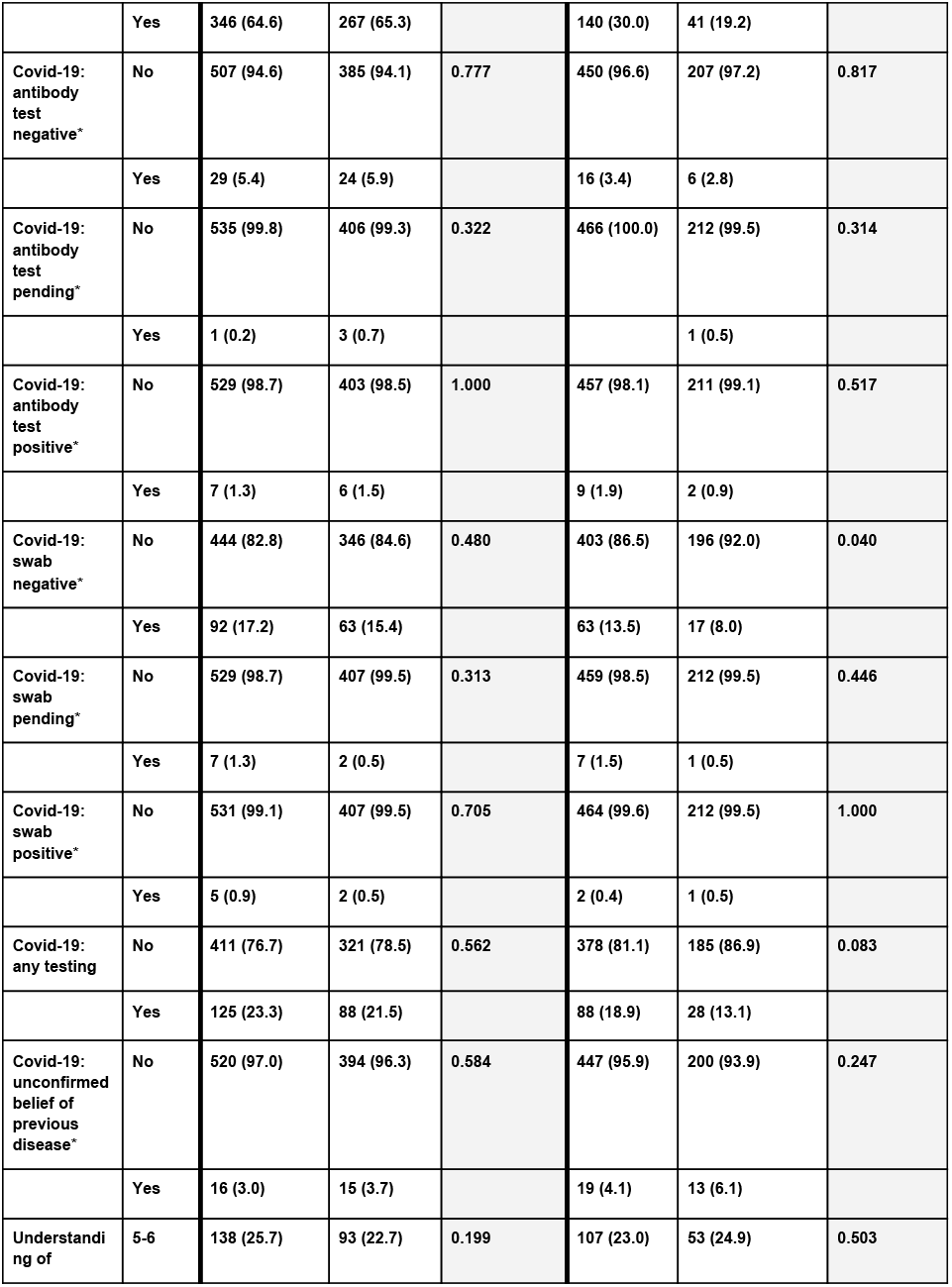
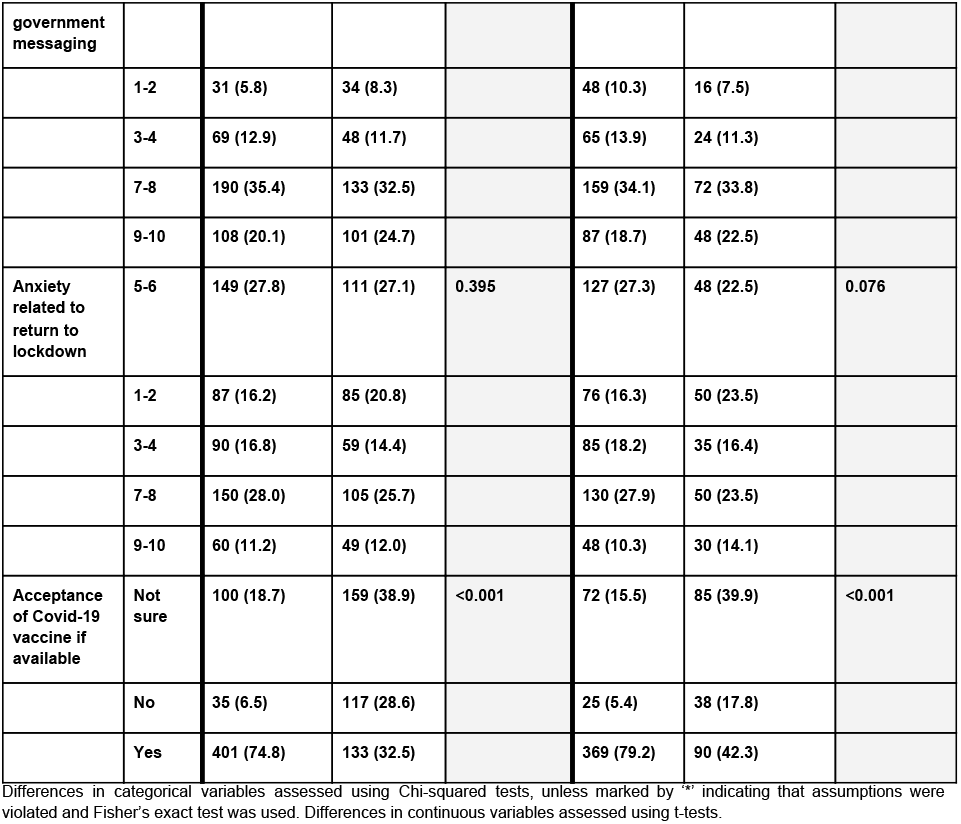
Baseline demographics of the study population, grouped according to eligibility and acceptance of the flu vaccination in 2020/21

Responses to items in prior questionnaires in the series were used to complete information on participant ethnicity, additional vaccine eligibility criteria (including chronic disease), index of multiple deprivation (IMD) quintile (obtained from participant postcode), any healthcare utilisation since beginning of lockdown, whether the participant considered themselves at high risk from Covid-19, experience of any Covid-19 symptoms, self-reported understanding of government advice, anxiety related to a return to lockdown, and whether the participant would agree to receive a Covid-19 vaccine if available.

### Inclusion and Exclusion Criteria

All participants were aged 18 or above. Participants must have answered questionnaires in weeks 9, 10, 13 and 16 to be included in the analysis, and answered ‘no’ to a question assessing whether they routinely received flu vaccination. Participants submitting incomplete or inconsistent responses to the questions on flu vaccination were excluded, as were those who answered ‘prefer not to say’ for ethnicity and who were missing responses for other variables required in the analysis, with the exception of post-code. Responses submitted later than four days from the time of the questionnaire launch were excluded.

### Data analysis

Age was categorised into 10 year age bands between 18-29 and 70+ to allow for easier interpretation of a potential non-linear relationship between age and responses to flu vaccination. The 10-point scale measurements of ‘anxiety related to return to lockdown’ and ‘understanding of government messaging’ were re-grouped into categories of 1-2, 3-4, 5-6, 7-8, and 9-10, and ethnicity was categorised into five groups due to low numbers in some categories. Descriptive statistics are reported for the dataset broken down according to study groups (described below). Differences in categorical variables were assessed by chi-square tests or Fisher’s exact test where chi-square test assumptions were violated, and differences in continuous variables were assessed using t-tests. P values <0.05 were considered statistically significant.

The effect of variables of interest on inclination to receive a flu vaccination were calculated using univariate and multivariate logistic regression models. The relationship between age (the only continuous variable) and the log-odds of receipt of flu vaccination were plotted and visually inspected. If the effect appeared linear, age was included as a linear variable, otherwise it was included as a categorical variable. All data were analysed in R version 3.6.2. Variables with low numbers in categories were not included in the multivariable analyses. ‘Acceptance of Covid-19 vaccine if available’ was deemed likely to be highly correlated with ‘accepting flu vaccine in 2020/21’ and was not included in multivariable models to allow greater interpretation of other predictors. Multi-collinearity was assessed by calculation of the variance inflation factor (VIF), with variables with a VIF >5 (indicating substantial multicollinearity) removed from the model.

#### Definition of Study Groups

The analyses in this study were confined to those participants eligible for free NHS flu vaccination in 2020/21 who indicated they had previously not routinely received it (this group is the greatest unknown for planning resourcing and targeting public health campaigns to maximise uptake). This previously unvaccinated group were either previously eligible (main criteria up to 2019/20 were age over 65, eligible comorbidity and working in the healthcare sector),^19^ or newly eligible for the expanded 2020/21 programme (age over 50). Further stratification according to whether or not the flu vaccine would be accepted in 2020/21 generated four groups: 1.) Previously eligible, newly responding ‘yes’, 2.) Previously eligible, still responding ‘no’, 3.) Newly eligible, responding ‘yes’, and 4.) Newly eligible, responding ‘no’.

#### Potential Change from Previously Receiving Vaccine to Declining in 2020/21

Without assuming those vaccinated last year would continue this habit, a specific question was posed in order to also measure if any routinely vaccinated participants would not do so again in 2020/21.

#### Households with Schoolchildren

Responses from participants in the four groups regarding presence of school age children in their household are reported and, if offered, whether they would want any of these children to receive flu vaccination in 2020/21.

#### Network Analysis

Each participant previously not receiving flu vaccination was asked to qualify their yes/no response to whether they would accept it in 2020/21 using a free text response option. Three researchers, blinded to the response on vaccine acceptance, each independently coded the content of 100 responses according to prospectively identified themes. A consensus was then reached to define the main themes for coding the remaining responses. Multiple themes could be coded for within a response. For example, “I will have the flu vaccine this year because I want to protect myself, and my GP says it’s a good idea” would count as “precaution for myself”, “GP” and “advice.” Similarly, “I don’t think I need it because I’ve never had the flu before” would count as “unnecessary” and “no previous flu.”

A network analysis,^20,21^ was generated for each one of the four groups using the Networkx package in Python (version 3.7). Dimensions of centrality and overall topography of nodes was not applicable; the network could therefore be laid out in a comprehensible circular ‘shell’ arrangement. Each network was limited to the ten most represented themes within each group’s responses. The size of the nodes corresponded to the frequency of the theme. Thickness of the lines (edges) between nodes represented the frequency of co-occurrence of those themes within responses. Nodes were colour coded to reflect positive, negative and neutral sentiment of themes.

Separately, reasons for healthworker’s continued non-vaccination in 2020/21 were reported descriptively. A full list of the themes with examples is available in table 2 of the supplementary materials table 2.

## RESULTS

6,641 respondents aged ≥18 completed the week 16 questionnaire on flu vaccination in the predefined time period, as well as the requisite previous weeks needed to complete baseline characteristics. Of these, 208 (3.1%) were missing answers to one or more essential variables and were removed, to leave 6,433 complete responses. Exclusion of those who did not meet the new or old eligibility criteria for the flu vaccine by age, comorbidity status, and/or health worker status (N=769) left 5,664 participants, of whom 4,641 (81.9%) were ‘previously eligible’ and 1,023 (18.1%) were ‘newly eligible’ participants. Finally, we excluded those who stated that they had routinely received the flu vaccination among ‘previously eligible’ participants (N=3,696, 79.6%) and among ‘newly eligible’ participants (N=344, 33.6%). The total number of previously (N=945) and newly eligible (N=679) unvaccinated participants was 1,624.

Of the previously eligible participants, those who had previously declined the vaccine were more likely to be younger (median age 61 years, IQR 51 to 67 vs median age 67 years, IQR 58 to 73), female (55.0% vs 46.7%), have chronic neurological disease (10.8% vs 6.5%), work in the health sector (10.2% vs 7.8%) and be in a lower IMD quintile, and were less likely to have chronic respiratory disease (14.5% vs 20.5%) and chronic heart disease (7.0% vs 12.2%) than those receiving the vaccine.

Of the newly eligible participants, when compared with those who had received the vaccine despite being ineligible by NHS criteria, those who had not received the vaccine were more likely to be younger (mean age 57, IQR 54 to 61 vs median age 59, IQR 55 to 63) and in a lower IMD quintile. Full description of differences is available in Supplementary Table 3. 309 (4.5%) of all respondents who indicated having received the flu vaccine in 2019/20 responded they did not intend to repeat this in 2020/21.

### Change in Acceptance and Uptake of flu Vaccine in 2020/2021

Summary statistics for groups broken down according to vaccine eligibility and acceptance of the flu vaccine in 2020/21 are shown in Table 1. 536 (56.7%) of those previously eligible but routinely not vaccinated intend to be vaccinated in 2020/21, increasing the vaccination rate in the entire previously eligible cohort from 79.6% to 91.2%. In contrast, 466 (68.6%) of the newly eligible reported they would accept vaccination in 2020/21

#### Univariate and Multivariable Analysis

Results from the univariate and multivariate analyses are displayed in Table 2 and Table 3. In the univariate analysis, acceptance of a Covid-19 vaccine if available was associated with acceptance of flu vaccination in 2020/21 in both groups compared to those who were unsure (OR = 4.79, 95%CI 3.50 to 6.61, OR = 4.84, 95% CI 3.29 to 7.17). 74.8% and 79.2% of those who would newly accept the flu vaccination who were previously eligible and newly eligible responded they would accept a Covid-19 vaccination, compared to 32.5% and 42.3% of those declining the flu vaccine.

In those who were previously eligible, answering ‘no’ in response to receiving a Covid-19 vaccination if offered was associated with a lower likelihood of wanting to receive the flu vaccination in 2020/21 (OR = 0.48, 95% CI 0.30 to 0.74), as was having a chronic neurological disease (OR = 0.65, 95% CI 0.43 to 0.98). Whilst those aged 60-69 were more likely to respond ‘yes’ than those aged 70+, (OR 1.48, 95% CI 1.02 to 2.14) there was no clear effect of age found in those below the age of 60. The multivariable analysis resulted in few substantial changes to effect estimates, with the exception of age, for which all estimates shifted upwards (showing a stronger association with an increased likelihood of answering ‘yes’ after adjustment for other variables).

In those who became newly eligible to receive the flu vaccine, there was an association between increased age (OR for 1-year increase in age = 1.07, 95% CI 1.03 to 1.12), IMD quintile, and considering oneself at high risk from Covid-19 (OR = 1.80, 95% CI 1.22 to 2.70) and answering ‘yes’ to receiving the flu vaccine if offered. Females were less likely to answer ‘yes’ (OR = 0.56, 95% CI 0.40 to 0.79), as were those who rated their anxiety about the lifting of lockdown as 1-2 (low anxiety) (OR = 0.57, 95% CI 0.35 to 0.93, compared to those rating it 5-6). The multivariable analysis resulted in minimal changes to the estimates.

#### Subgroup Analysis of Households with Schoolchildren

1419 (87.4%) answered the question items pertaining to flu vaccination of school children. Among these, 150 responded they had school children in their household and answered ‘yes’ or ‘no’ to whether they would want any children to be vaccinated in 2020/21 if offered. Among the 71 participants who were previously eligible but not routinely vaccinated, 33/40 (82.5%) of those who would accept vaccination in 2020/21 would also vaccinate children, compared to 8/31 (25.8%) of those who would not accept the flu vaccine for themselves (Fisher’s exact test p<0.001).

Among the 79 participants who were previously unvaccinated and newly eligible in 2020/21, 46 (82.1%) of those who would get a flu vaccine this year would want their child to have it also, compared to 10 (43.5%) of those who would not get the flu vaccine for themselves (Fisher’s exact test p = 0.001).

#### Subgroup Analysis of Healthcare Workers

49 (51.0%) of previously unvaccinated healthcare workers would accept the vaccine in 2020/21, compared to 47 (49.9%) who would continue to decline.

### Network Analysis

A free text response qualifying why participants would/would not accept flu vaccination in 2020/21 was submitted by 834 (88.3%) from the previously eligible, unvaccinated and 619 (91.2%) of the newly eligible group. These were coded according to 45 themes (full list in supplementary table 2). Figure 4 displays network diagrams for the ten most common themes for each group.

**Figure 1.**
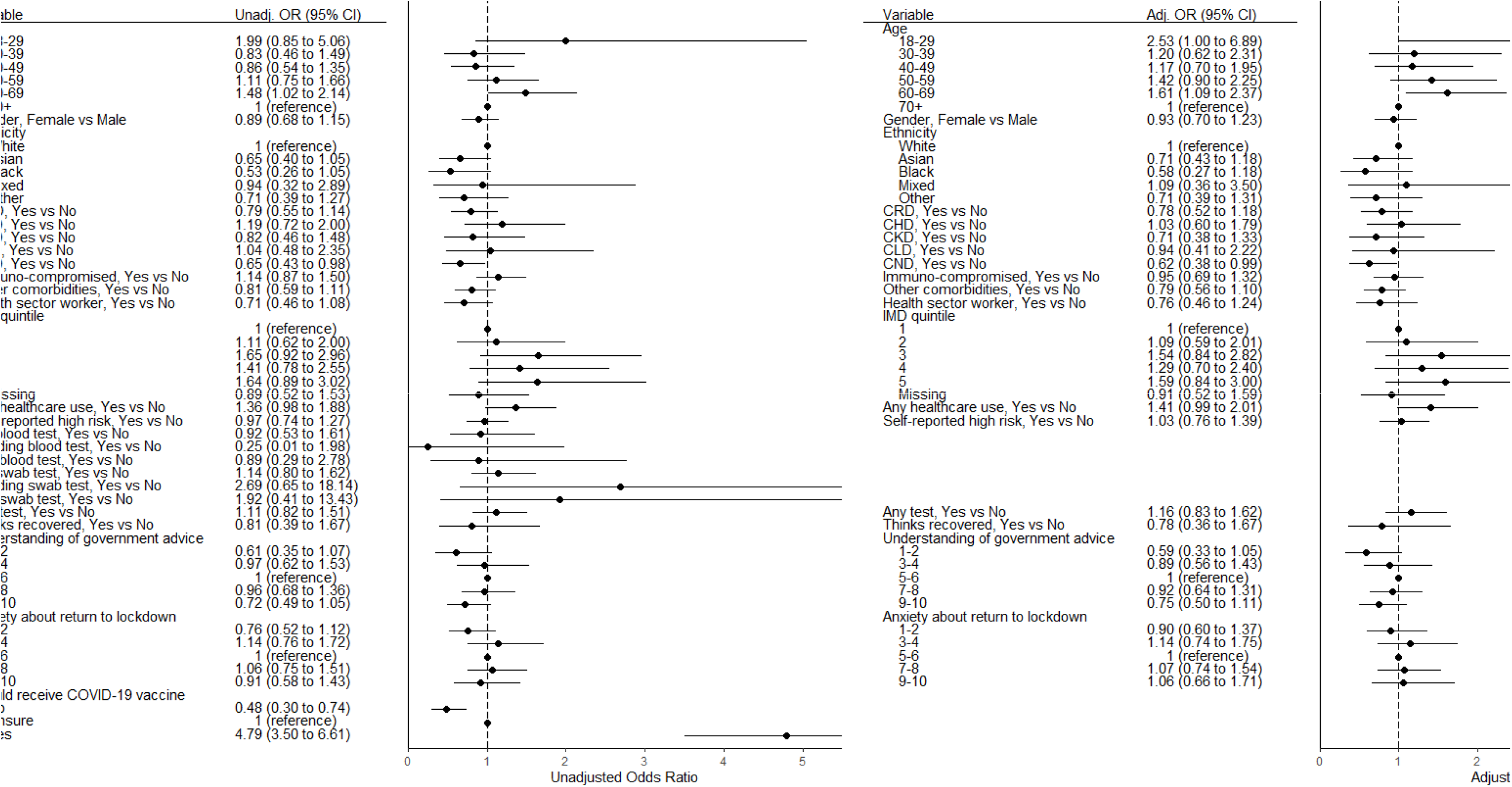
Participant inclusion flow diagram based on responses to questionnaires capturing variables required for analysis.

**Figure 2.**
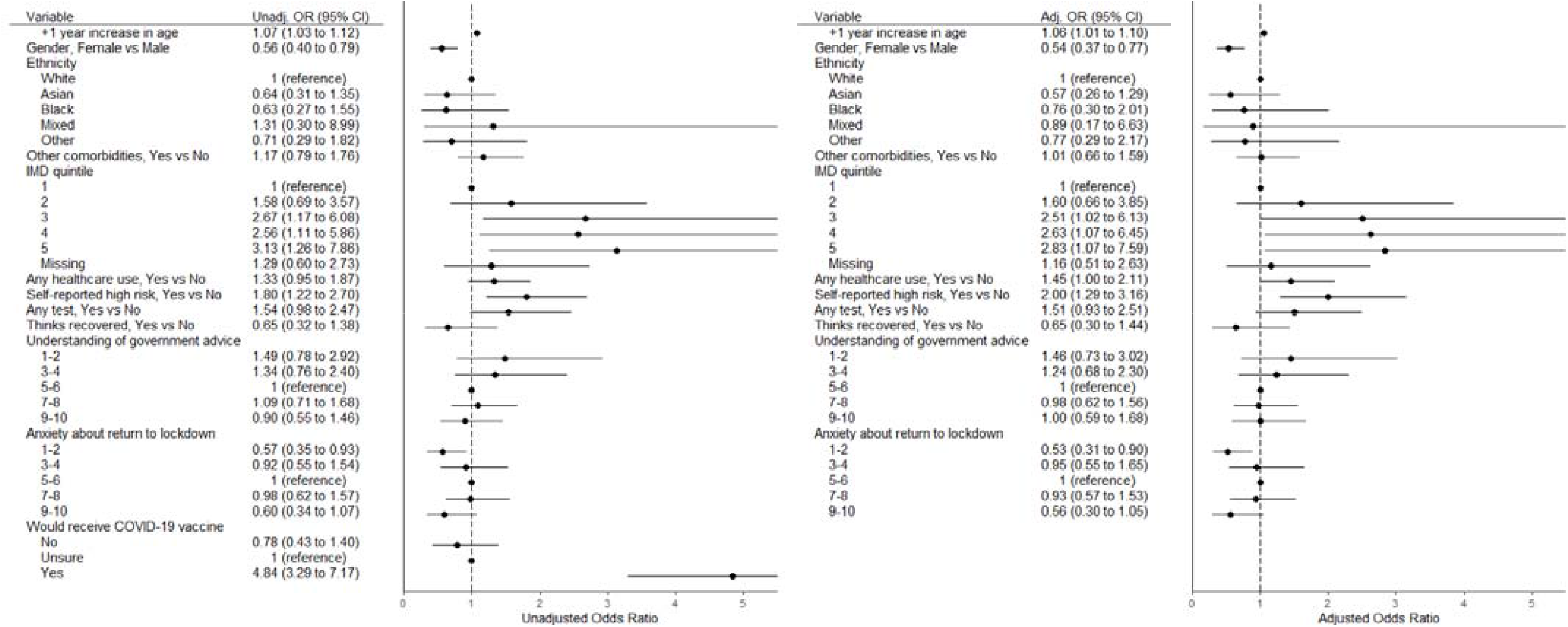
Unadjusted and adjusted logistic regression for associations with a ‘yes’ response in participants who would accept a flu vaccine in 2020/21 in those who were previously eligible but did not routinely receive flu vaccination. Adjusted odds ratio adjusted for every other variable in model (age, sex, ethnicity, disease, IMD quintile, health care utilisation, considering oneself at high risk for Covid-19, undertaking any Covid-19 test, believing oneself to have had Covid-19, understanding of government advice, anxiety related to a return to lockdown).

**Figure 3.**
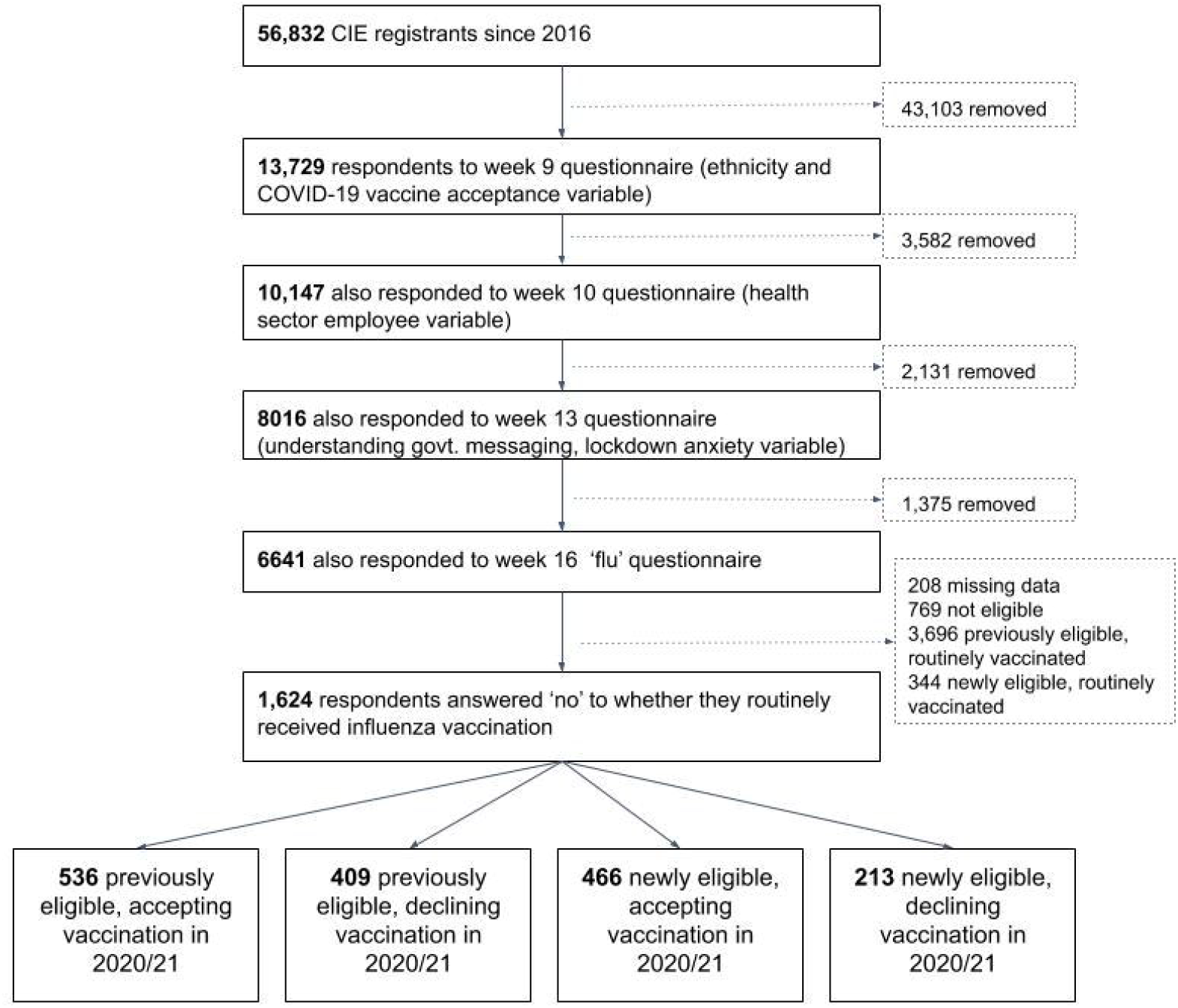
Unadjusted and adjusted logistic regression for associations with a ‘yes’ response in participants who would accept a flu vaccine in 2020/21 in those who were newly eligible and did not routinely receive flu vaccination already. Adjusted odds ratio adjusted for every other variable in model (age, sex, ethnicity, disease, IMD quintile, health care utilisation, considering oneself at high risk for Covid-19, undertaking any Covid-19 test, believing oneself to have had Covid-19, understanding of government advice, anxiety related to a return to lockdown).

**Figure 4.**
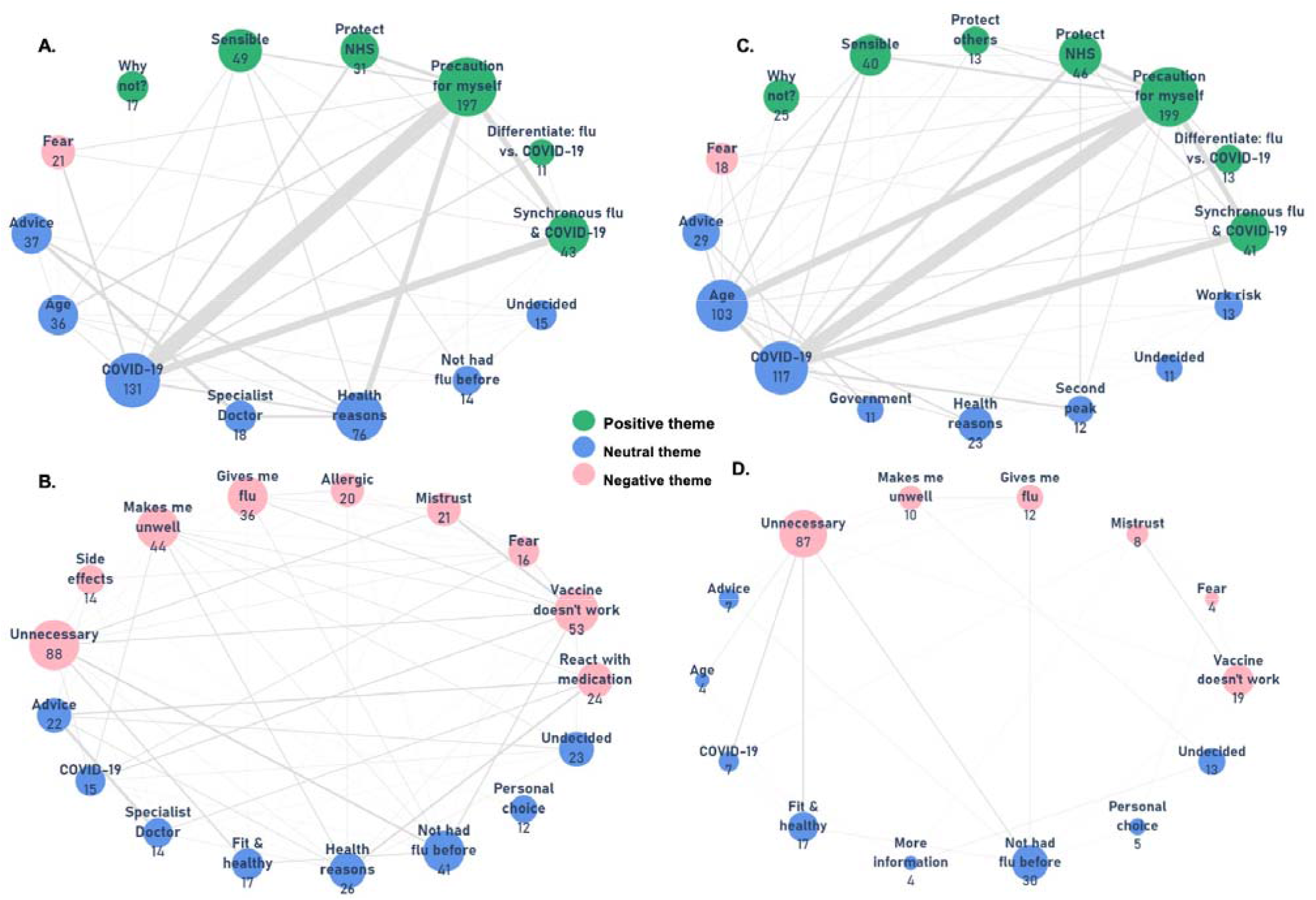
Network analysis of free-text responses from previously eligible respondents who had previously not accepted the flu vaccine but would accept it in 2020/21 (A; n = 478), or continue to decline (B; n = 356); responses from newly eligible participants who would accept vaccination (C; n = 432) or decline (D; n = 187). Colour-coded sentiment of nodes where green = positive, red = negative, blue = neutral. A connecting line between nodes implies at least one response where themes of connected nodes co-occurred; the thickness of the line corresponds to the frequency of co-occurrence.

#### Previously Eligible, Newly Accepting flu Vaccination in 2020/21

478 (89.8%) responses were coded. The three most frequent themes were ‘precaution for myself’ 197 (41.2%), ‘Covid-19’ 131 (27.4%) and ‘health reasons’ 76 (15.9%). Among responses containing ‘precaution for myself’, 71 (36.0%) qualified this with ‘Covid-19’, 26 (13.2%) with ‘synchronous flu & Covid-19’ and 26 (13.2%) with ‘health reasons’.

#### Previously Eligible, Continuing to Decline flu Vaccination in 2020/21

356 (87.0%) responses were coded. The three most frequent themes were ‘unnecessary’ (88, 24.7%), ‘vaccine doesn’t work’ (53, 14.9%), and ‘makes me unwell’ (44, 12.4%). Responses were more discrete than in those newly accepting vaccination. Among responses containing ‘Unnecessary’, 9 (10.2%) said this was due to ‘no previous flu’ and 7 (8.0%) due to being ‘fit and healthy’.

#### Newly Eligible, Accepting flu Vaccination in 2020/21

432 (92.7%) responses were coded. The three most frequent themes were ‘precaution for myself’ (199, 46.1%), ‘Covid-19’ (117, 27.1%) and ‘age’ (103, 23.9%). Among responses containing ‘precaution for myself’, 58 (29.1%) qualified this with ‘Covid-19’, 36 (18.1%) with ‘age’ and 24 (12.1%) with ‘synchronous flu & Covid-19’.

#### Newly Eligible, Declining flu Vaccination in 2020/21

187 (87.8%) responses were coded. The three most frequent themes were ‘unnecessary’ (87, 46.5%), ‘not had flu before’ (30, 16.0%) and ‘vaccine doesn’t work’ (19, 10.2%). Responses were again more discrete than in those accepting vaccination. Where ‘unnecessary’ was qualified, it was most commonly with ‘fit and healthy’ (7, 8.0%)

#### Subgroup analysis for healthcare worker continued non-vaccination

89 (85.6%) healthcare workers reporting previous non-vaccination submitted qualifying responses, among whom 47 were from those newly accepting and 42 continuing to decline in 2020/21. For the former, ‘precaution for myself’ (17, 36.2%), ‘Covid-19’ (16, 34.0%) and ‘health reasons’ (8, 17.0%) were most cited. In those continuing to decline, most frequent reasons were ‘gives me flu’ (10, 23.8%), ‘vaccine doesn’t work’ (8, 19.0%) and ‘unnecessary’ (6, 14.3%).

## DISCUSSION

In this UK-wide survey of 6,641 participants, we confined analysis of our questionnaire measuring planned uptake of flu vaccination in 2020/21 to 945 previously eligible but unvaccinated participants and 679 participants newly eligible by the expanded criteria for free NHS flu vaccination. 56.7% of previously eligible but unvaccinated participants now intend to receive the flu vaccine in 2020/21 (increasing the vaccination rate in the entire previously eligible cohort from 79.6% to 91.2%). Among the newly eligible, 68.6% would accept vaccination. On the basis of these findings, of the estimated 32.4 million (48.8%) of the UK population eligible for influenza vaccination this year 26 million (80%) are planning to have it.

Among participants with school age children, of those previously eligible who now intend to be vaccinated themselves, 82.5% also intend to vaccinate their children in 2020/21 compared to 25.8% of those who would still not accept the influenza vaccine for themselves. Among the newly eligible respondents this was 82.1% and 43.5%, respectively. 49.9% of the previously unvaccinated healthcare workers would continue to decline the vaccine in 2020/21.

Those previously eligible aged 60-69 were 48% more likely to respond ‘yes’ to vaccination in 2020/21 than those aged 70+ (who as the group highest at-risk from Covid-19 may be less inclined to risk exposure to get a flu vaccine); there was no clear effect of age in those under 60. The effect of chronic neurological disease (45% less likely) may be explained by patients receiving specific therapy (such as for multiple sclerosis) contraindicating flu vaccination.

In the newly eligible, increased age showed a positive association (OR for 1-year increase in age = 1.07, 95% CI 1.03 to 1.12); those with low levels of anxiety around lifting of lockdown were 43% less likely to accept vaccination than those with moderate levels. Considering oneself at high risk from Covid-19 was associated with an 80% increase in accepting vaccination in 2020/21. In both groups, the effect of IMD showed that declining vaccination follows a social gradient.

Network analysis of responses from previously eligible unvaccinated and newly eligible participants accepting vaccination in 2020/21 showed that the most frequent themes were ‘precaution for myself’ (41.2% and 46.1%) and ‘Covid-19’ (27.4% and 27.1%), where the former was qualified by the latter in 36% and 29.1% of responses in each group, respectively. Among the previously and newly eligible not accepting vaccination, collectively, misinformed themes of ‘makes me unwell’, ‘gives me flu’ and ‘vaccine doesn’t work’ were present across 37.4% and 21.9% of responses, respectively.

The reasons for previously not receiving vaccination were not influenced by the context of the Covid-19 pandemic and therefore have long-term relevance. Public trust is critical for vaccine confidence,^22,23^ which must be underpinned by clear messaging campaigns. Once formed, a false belief that the flu vaccine ‘gives me the flu’ is difficult to correct.^24^ The nearly 11 million newly eligible, as manifest in this study, harbour fewer false vaccine preconceptions than those previously eligible. This highlights an opportunity to consolidate positive attitudes towards vaccination.

Emergence from the Covid-19 pandemic hinges on a vaccine,^25^ therefore successful new approaches to improving uptake of established vaccination programmes could inform wider implementation strategies. A recent survey of 2,700 participants suggested only 30% would be willing to be in the first cohort to receive an approved Covid-19 vaccine.^26^ However, there is some suggestion that broadly, enthusiasm for vaccination increases during a pandemic, prior to, and immediately following the release of a novel vaccine.^27^ This study highlights that circulation of one pandemic virus fuels uptake for vaccination against another.

The current pandemic takes place at a time of unprecedented access to information and education on respiratory viruses and vaccine development, in a UK population with recently increased trust in scientific reporting.^28,29^ Nonetheless, and more so than in previous pandemics, social media is proving particularly damaging by hosting a proliferation of misinformation.^30^ Transparency in how a vaccine is being developed must be accompanied by assurances that safety and efficacy are critical -- rushing through a subsequently problematic vaccine could harm public trust long-term.^31^

In spite of the fact 50% of healthcare professionals who refused flu vaccine previously still do not intend to have it, this study suggests that the UK population feels a sense of duty to the NHS; 8.5% of those newly accepting vaccination cited ‘protect the NHS’ as their reason. Such messaging, as was used to encourage adherence to the government’s ‘stay at home’ policy during the height of the first wave of the pandemic,^32^ could also be leveraged to increase uptake of flu and more ambitious vaccination programmes: the UK government is exploring the possibility of co-administering a flu-Covid-19 vaccination.^33^ There is some suggestion that flu vaccination can itself impact Covid-19 rates of disease acquisition, transition and ameliorate symptoms. ^34^

Public Health England’s finding that co-infection doubles the risk of death^13^ was publicised after collection of the data described in this study; nonetheless, our results indicate that specifically avoidance of ‘synchronous flu and Covid-19’ (15%) and ‘differentiating flu vs. Covid-19’ (4.5%) are motivators for new flu vaccine uptake for the 2020/21 season. This suggests that the UK public already perceived the risk from a convergence of both viruses.

This study has several limitations. These results are only indicative; whether participants stick to their response when faced with flu vaccination is uncertain. However, previous studies have shown good correlation between declared questionnaire responses and subsequent behaviour.^35–37^Those who turned 65 or became a healthcare worker in the last year and who did not receive a flu vaccination in 2019/20 were potentially mis-classified. Use of the CIE implies a higher agency over one’s health, and notably the previously eligible population had a higher baseline uptake (79.6%) than last year’s national average (70.6%).

The number of healthcare workers in the sample was low, limiting the generalisability of reasons for and against new uptake, though the prevalent belief that the vaccine ‘gives me flu’ is reflected in other studies of healthcare workers,^38^ among whom low uptake remains a challenge.^39^ Misconceptions and false beliefs around the flu vaccine are common and harmful, and follow a socioeconomic gradient.^40^ Therefore a further limitation is the under-representation of ethnic minorities, also known to have lower uptake of all forms of vaccination.^41^ As well as potentially explaining the relative absence in the network analysis of logistical reasons, such as getting time off work to attend an appointment, this shortcoming in representativeness may have resulted in slightly overestimated acceptance rates. The need to understand the Covid-19 pandemic’s impact on acceptability of a widened flu vaccination programme was time-sensitive, prohibiting the generation of question items using, for example, in-depth Delphi methods and full psychometric evaluation of validity.

This study used a semiquantitative approach with content classification of rich free-text responses for network analysis, avoiding the biases of pre-defined options for participants to choose from. However, posing specific questions on socioeconomic, cultural, sociopolitical and religious influence on flu vaccination may have identified further determinants that could be influenced by focused health policy messaging.

Lastly, it is important to note that even in the potential presence of a virulent, highly contagious strain of seasonal influenza, winter 2020/21 will see ongoing widespread measures to contain Covid-19 that, if maintained, will likely also result in declining transmission rates of other respiratory viruses, with rates driven down even further if flu vaccine uptake is as high as this study suggests.

## Conclusion

The Covid-19 pandemic has increased acceptance of flu vaccination in 2020/21 from 79.6% to 91.2% in those previously eligible, and for the 69% of those newly eligible. This high anticipated vaccination rate (to 26 million (80%) of the UK population) requires appropriate planning, but can be further increased with effective messaging campaigns to address negative misconceptions about flu vaccination, which may also help prepare for future Covid-19 vaccination. It remains of concern that 50% of healthcare professionals who refused it previously still do not intend to have the flu vaccine, but emerging additional evidence for benefit during Covid-19 may impact this. Maximising influenza vaccination requires informed planning of vaccine supply and public health messaging if failure once again to meet the demands of a public health imperative is to be avoided.

## Data Availability

Imperial College Healthcare NHS Trust is the data controller. The datasets analysed in this study are not publicly available but can be shared for scientific collaboration subject to meeting requirements of the institution's data protection policy.

## Author Contribution

Patrik Bachtiger: study design, data collection, literature review, data analysis, figures, writing

Alexander Adamson: study design, literature review, figures, data analysis, writing

Ji-jian Chow: figures, data analysis, writing Rupa Sisodia: figures, data analysis, writing

Jennifer K Quint: study design, literature review, data analysis, figures, writing

Nicholas S Peters: study design, data collection, literature review, data analysis, figures, writing

NSP is the guarantor. The corresponding author attests that all listed authors meet authorship criteria and that no others meeting the criteria have been omitted.

The lead author affirms that this manuscript is an honest, accurate, and transparent account of the study being reported; that no important aspects of the study have been omitted; and that any discrepancies from the study as planned (and, if relevant, registered) have been explained.

## Ethical approval

The weekly questionnaire was a direct care tool for patients to self-monitor their wellbeing during the Covid-19 pandemic. Review by the Imperial College Healthcare NHS Trust Data Protection Office advised ethical approval for data analysis and publication was not required. Participants were informed prior to completing responses that these would be anonymised to inform local and national health policy and were free to opt out.

## Dissemination to participants and related patient and public communities

We plan to disseminate these findings to participants in our Trust’s annual online newsletter.

## Acknowledgements

We acknowledge the combined inputs from Imperial College London colleagues expert in questionnaire design and analysis from our departments including Faculty of Medicine, School of Public Health and Institute of Global Health Innovation.

