## supplementary for "The Impact of the Covid-19 Pandemic on Uptake of Influenza Vaccine: A UK-Wide Observational Study"

**A UK-Wide Observational Study”, September 2020**

**Supplementary Materials**

**Figure 1.** Map of UK showing CIE registrants by postcode (each blue dot indicates at least one registrant in that postcode areas).

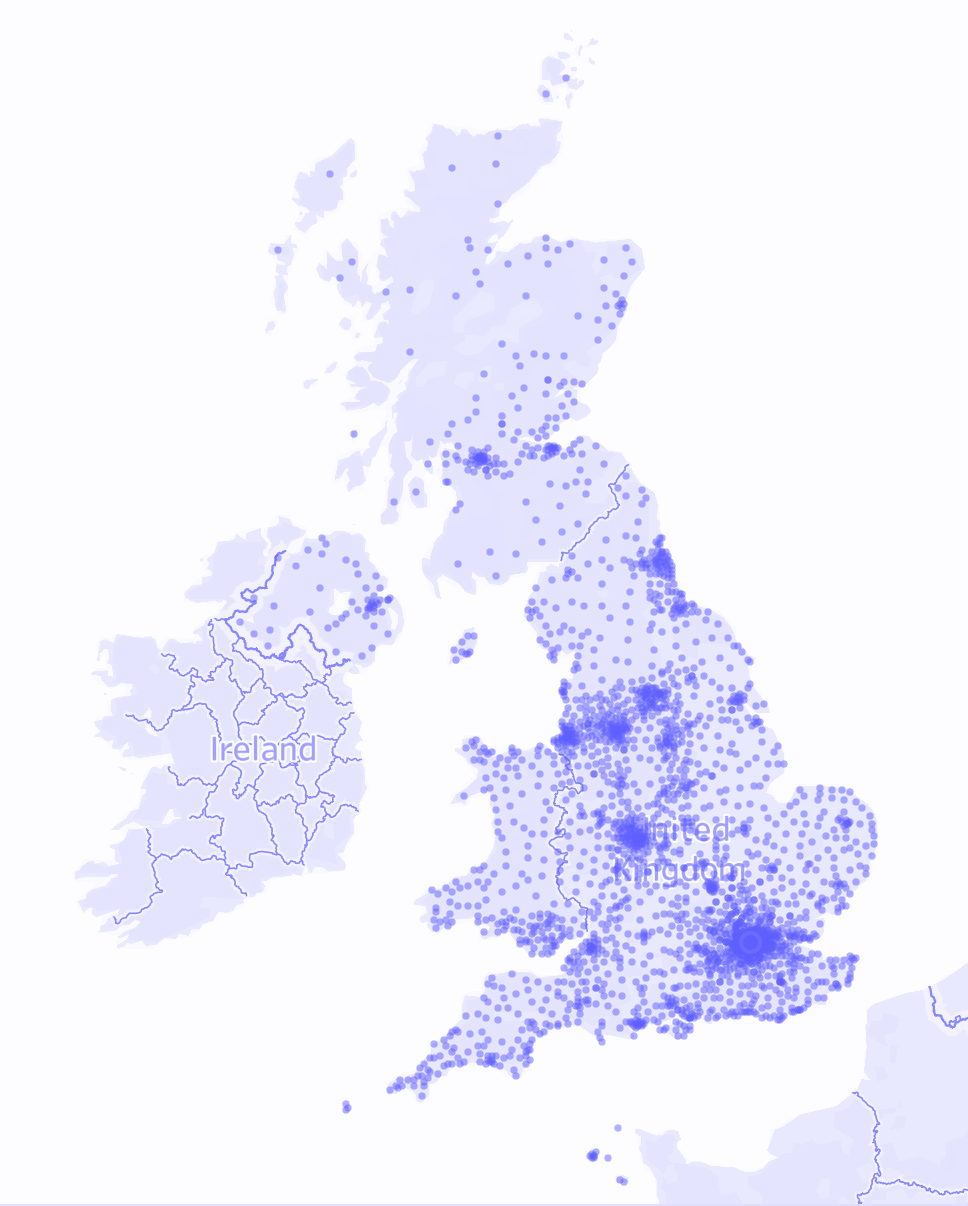

**Table 1:** Imperial College Healthcare NHS Trust weekly self-monitored wellbeing questionnaire for patients (week 16 items).

**Core questions:**

**How do you feel physically today?**

1 = worst … 10 = best

**How is your mood today?**

1 = worst … 10 = best

**Have you routinely had the flu jab in recent years?**

Yes - because of my age and/or medical conditions

Yes - other reason [for health workers etc]

No - but I should because of my age or medical conditions

No - not at high risk from flu

**If you do NOT usually have the flu jab would you have it this year if offered?**

I usually have it anyway

Yes

No

**If you answered YES or NO to the question above please give your reasons:**

[FREE TEXT]

**In the last week, have you had a cough?**

No

Yes -- I have a new, dry cough

Yes -- I have a new cough bringing up phlegm

Yes -- but I usually have a cough like this

Yes -- my usual cough has worsened

**In the last week, have you experienced unusual shortness of breath compared to what's normal for you?**

No

Yes -- Mild symptoms, slight shortness of breath during ordinary activity

Yes – Significant symptoms, breathing is comfortable only at rest

Yes -- Severe symptoms, breathing difficult even at rest

**In the last week, have you had a fever and did you take your temperature?**

I have NOT felt feverish

I have felt feverish but did not check my temperature

I felt feverish and my temperature was BELOW 38 degrees Celcius

I felt feverish and my temperature measured ABOVE 38 degrees Celcius

**In the last week, have you experienced any of these other symptoms? Please tick all that apply**

I haven't had any new symptoms

Loss of sense of smell

Loss of appetite (skipping meals)

Diarrhoea

Vomiting

Fatigue

Sneezing

Chest pain / tightness

Sore throat

Runny nose

Itchy eyes

Headache

Joint pain / aches

Rash

**In the last week, has anyone in your household had a new cough or fever?**

Not applicable

No

Yes

**Have you or anyone in your house been tested for coronavirus? Please tick all that apply**

No testing

I have not been tested – BUT I think I have already had coronavirus and recovered

I was tested with a SWAB -- positive result

I was tested with a SWAB -- awaiting result

I was tested with a SWAB -- negative result	Household member tested with a SWAB -- positive result

Household member tested with a SWAB -- awaiting result

Household member tested with a SWAB -- negative result

I was tested with a ANTIBODY blood test-- positive result

I was tested with a ANTIBODY blood test -- awaiting result

I was tested with a ANTIBODY blood test -- negative result

Household member tested with a ANTIBODY blood test -- positive result

Household member tested with a ANTIBODY blood test -- awaiting result	Household member tested with a ANTIBODY blood test -- negative result

**Have you had any healthcare contact since the lockdown started? Please tick all that apply**

No

I have had appointment(s) cancelled and not yet rescheduled

Yes - remote appointment with my GP (phone/video)

Yes - I attended my GP practice for an appointment

Yes - I have had new contact with mental health services (remote or in person, including counselling)

Yes - remote appointment with hospital (phone/video)

Yes - I attended hospital for an appointment

Yes - attended Accident and Emergency	Yes -- I was admitted to hospital (not because of coronavirus)

Yes -- I was admitted to hospital with symptoms of coronavirus

**Table 2:** Full list of themes from free text responses for or against receiving vaccination in 2020/21.

| **Node Text** | **Example** |
| --- | --- |
| **Advice** | "Advised to get it", "Told I should have it", "Advice from ..." |
| **Affordability** | "I couldn't afford to pay for it", "If I don't have to pay I'll have it" |
| **Age** | "Age", "Because of my age", "Because I'm getting old" |
| **Allergic** | "I'm allergic to the vaccine", "I'm allergic to eggs", "I'm worried about allergies" |
| **Boost immunity** | "Boost my immune system", "Strengthen my immune system" |
| **Covid-19** | "Covid-19", "covid", "coronavirus" |
| **Differentiate: flu vs. Covid-19** | "If I get Covid-19 at least I'll know it's probably not flu", "That way I'll know if it's covid" |
| **Discrimination** | "I will be discriminated against if I don't have it" |
| **Family/friends unwell from vaccine** | "My mother was ill for weeks after the flu jab", "My son was very poorly afterwards" |
| **Fear** | "Fear", "Afraid", "Scared" |
| **Fit & healthy** | "I'm fit and healthy", "I'm in good health", "I never get sick" |
| **Gives me flu** | "The vaccine is a dose of flu", "I had the flu jab years ago and it gave me the worst flu ever", "I got sick with the flu after having the jab" |
| **Government** | "Government", "the authorities", "policymakers" |
| **GP** | "GP", "general practice" |
| **Health reasons** | "Health conditions", "my diabetes", "my heart disease", "my multiple sclerosis" |
| **Household member** | "My mother is immunocompromised", "I get it because my husband is at-risk" |
| **Logistics** | "I can't get appointments", "I can't travel to get one" |
| **Makes me unwell** | "The flu jab made me very ill", "I was ill for weeks after the flu jab", "I was in hospital for a week after getting the jab" |
| **Misbelief in vaccines** | "I don't believe vaccines are good for you", "I don't believe vaccines are the right treatment" |
| **Mistrust** | "Don't trust the flu jab", "Don't trust what's in the vaccine" |
| **More information** | "I need more information", "I don't understand why it's needed" |
| **Natural health** | "I prefer natural remedies", "I want to use my natural immunity to fight flu", "Alternative therapies are better for flu" |
| **Needle phobia** | "I hate needles", "I have a needle phobia" |
| **No Covid-19 protection** | "It won't' protect against covid", "Doesn't include strains against covid" |
| **Not had flu before** | "Not had flu before", "Never had flu before", "Not had flu in the past" |
| **Personal choice** | "Personal choice", "I prefer not to" |
| **Precaution for myself** | "Precaution", "to protect myself", "to keep me safe" |
| **Pregnancy** | "Pregnancy", "I'm pregnant" |
| **Prioritise others** | "Prioritise others", "Not enough supply so give it to those who need it first" |
| **Protect NHS** | "Protect the NHS", "relieve pressure on the NHS", "support the health system" |
| **Protect others** | "Protect others", "keep others safe" |
| **React with medication** | "Reacts with my medication" |
| **Responsibility** | "It's my duty" |
| **Risk of exposure** | "I don't want to go to the GP to get a flu jab and risk getting Covid-19" |
| **Second peak** | "Second peak", "second wave", "second surge" |
| **Sensible** | "Sensible", "Good idea" |
| **Side effects** | "Potential side effects" |
| **Specialist Doctor** | "My specialist doctor", "my consultant", "nephrologist", "rheumatologist" |
| **Synchronous flu & Covid-19** | "I don't want to get flu and Covid-19 at the same time" |
| **Undecided** | "Not sure actually", "I'm not completely sure" |
| **Unnecessary** | "Unnecessary", "Don't need it", "No point" |
| **Vaccine doesn't work** | "The vaccine is ineffective", "The vaccine never has the right strains", "The flu jab doesn't work" |
| **Why not?** | "Why not?" |
| **Work provides** | "Available at workplace", "employer offers it" |
| **Work risk** | "I'm at risk due to my work" |

**Table 3: Baseline characteristics of those who received the flu vaccination last year compare to those who did not receive it, broken down into previously eligible and newly eligible**

|  |  | **Previously eligible, did not have flu vaccine last year** | **Previously eligible, Did have flu vaccine last year** | **p** | **Newly eligible, did not have flu vaccine last year** | **Newly eligible, Did have flu vaccine last year** | **p** |
| --- | --- | --- | --- | --- | --- | --- | --- |
| **Total N (%)** |  | **945 (20.4)** | **3696 (79.6)** |  | **679 (66.4)** | **344 (33.6)** |  |
| **Age** | **Median (IQR)** | **61 (51 to 67)** | **67 (58 to 73)** | **<0.001** | **57 (54 to 61)** | **59 (55 to 63)** | **<0.001** |
| **Sex** | **Male** | **425 (45.0)** | **1969 (53.3)** | **<0.001** | **283 (41.7)** | **153 (44.5)** | **0.431** |
|  | **Female** | **520 (55.0)** | **1727 (46.7)** |  | **396 (58.3)** | **191 (55.5)** |  |
| **Ethnicity** | **White** | **773 (81.8)** | **3128 (84.6)** | **0.255** | **596 (87.8)** | **287 (83.4)** | **0.115** |
|  | **Asian** | **75 (7.9)** | **267 (7.2)** |  | **32 (4.7)** | **21 (6.1)** |  |
|  | **Black** | **35 (3.7)** | **103 (2.8)** |  | **22 (3.2)** | **13 (3.8)** |  |
|  | **Mixed** | **14 (1.5)** | **43 (1.2)** |  | **8 (1.2)** | **2 (0.6)** |  |
|  | **Other** | **48 (5.1)** | **155 (4.2)** |  | **21 (3.1)** | **21 (6.1)** |  |
| **Eligible disease** | **0** | **808 (85.5)** | **2939 (79.5)** | **<0.001** |  |  |  |
|  | **1** | **137 (14.5)** | **757 (20.5)** |  |  |  |  |
| **Chronic respiratory disease** | **No** | **879 (93.0)** | **3244 (87.8)** | **<0.001** |  |  |  |
|  | **Yes** | **66 (7.0)** | **452 (12.2)** |  |  |  |  |
| **Chronic heart disease** | **No** | **897 (94.9)** | **3457 (93.5)** | **0.133** |  |  |  |
|  | **Yes** | **48 (5.1)** | **239 (6.5)** |  |  |  |  |
| **Chronic kidney disease** | **No** | **919 (97.2)** | **3606 (97.6)** | **0.661** |  |  |  |
|  | **Yes** | **26 (2.8)** | **90 (2.4)** |  |  |  |  |
| **Chronic liver disease** | **No** | **843 (89.2)** | **3455 (93.5)** | **<0.001** |  |  |  |
|  | **Yes** | **102 (10.8)** | **241 (6.5)** |  |  |  |  |
| **Chronic neurological disease** | **No** | **612 (64.8)** | **2419 (65.4)** | **0.721** |  |  |  |
|  | **Yes** | **333 (35.2)** | **1277 (34.6)** |  |  |  |  |
| **Immunocompromised** | **No** | **749 (79.3)** | **2740 (74.1)** | **0.001** |  |  |  |
|  | **Yes** | **196 (20.7)** | **956 (25.9)** |  |  |  |  |
| **Other eligible co-morbidity** | **No** | **849 (89.8)** | **3409 (92.2)** | **0.020** |  |  |  |
|  | **Yes** | **96 (10.2)** | **287 (7.8)** |  |  |  |  |
| **Health sector employee** | **1** | **65 (6.9)** | **202 (5.5)** | **0.019** | **31 (4.6)** | **20 (5.8)** | **<0.001** |
|  | **2** | **142 (15.0)** | **515 (13.9)** |  | **105 (15.5)** | **42 (12.2)** |  |
| **IMD** | **3** | **166 (17.6)** | **635 (17.2)** |  | **123 (18.1)** | **36 (10.5)** |  |
|  | **4** | **140 (14.8)** | **674 (18.2)** |  | **115 (16.9)** | **73 (21.2)** |  |
|  | **5** | **123 (13.0)** | **571 (15.4)** |  | **72 (10.6)** | **61 (17.7)** |  |
|  | **(Missing)** | **309 (32.7)** | **1099 (29.7)** |  | **233 (34.3)** | **112 (32.6)** |  |
| **Healthcare utilisation** | **No** | **180 (19.0)** | **605 (16.4)** | **0.056** | **225 (33.1)** | **97 (28.2)** | **0.125** |
|  | **Yes** | **765 (81.0)** | **3091 (83.6)** |  | **454 (66.9)** | **247 (71.8)** |  |
| **Considering self at high risk from Covid-19** | **No** | **332 (35.1)** | **849 (23.0)** | **<0.001** | **498 (73.3)** | **207 (60.2)** | **<0.001** |
|  | **Yes** | **613 (64.9)** | **2847 (77.0)** |  | **181 (26.7)** | **137 (39.8)** |  |
| **Covid-19: antibody test negative*** | **No** | **892 (94.4)** | **3458 (93.6)** | **0.387** | **657 (96.8)** | **331 (96.2)** | **0.790** |
|  | **Yes** | **53 (5.6)** | **238 (6.4)** |  | **22 (3.2)** | **13 (3.8)** |  |
| **Covid-19: antibody test pending*** | **No** | **941 (99.6)** | **3675 (99.4)** | **0.769** | **678 (99.9)** | **341 (99.1)** | **0.221** |
|  | **Yes** | **4 (0.4)** | **21 (0.6)** |  | **1 (0.1)** | **3 (0.9)** |  |
| **Covid-19: antibody test positive*** | **No** | **932 (98.6)** | **3649 (98.7)** | **0.927** | **668 (98.4)** | **340 (98.8)** | **0.765** |
|  | **Yes** | **13 (1.4)** | **47 (1.3)** |  | **11 (1.6)** | **4 (1.2)** |  |
| **Covid-19: swab negative*** | **No** | **790 (83.6)** | **3114 (84.3)** | **0.658** | **599 (88.2)** | **297 (86.3)** | **0.446** |
|  | **Yes** | **155 (16.4)** | **582 (15.7)** |  | **80 (11.8)** | **47 (13.7)** |  |
| **Covid-19: swab pending*** | **No** | **936 (99.0)** | **3662 (99.1)** | **1.000** | **671 (98.8)** | **339 (98.5)** | **0.939** |
|  | **Yes** | **9 (1.0)** | **34 (0.9)** |  | **8 (1.2)** | **5 (1.5)** |  |
| **Covid-19: swab positive*** | **No** | **938 (99.3)** | **3677 (99.5)** | **0.556** | **676 (99.6)** | **342 (99.4)** | **1.000** |
|  | **Yes** | **7 (0.7)** | **19 (0.5)** |  | **3 (0.4)** | **2 (0.6)** |  |
| **Covid-19: any testing** | **No** | **732 (77.5)** | **2857 (77.3)** | **0.951** | **563 (82.9)** | **281 (81.7)** | **0.688** |
|  | **Yes** | **213 (22.5)** | **839 (22.7)** |  | **116 (17.1)** | **63 (18.3)** |  |
| **Covid-19: unconfirmed belief of previous disease*** | **No** | **914 (96.7)** | **3594 (97.2)** | **0.455** | **647 (95.3)** | **324 (94.2)** | **0.544** |
|  | **Yes** | **31 (3.3)** | **102 (2.8)** |  | **32 (4.7)** | **20 (5.8)** |  |
| **Understanding of government messaging** | **5-6** | **231 (24.4)** | **949 (25.7)** | **0.284** | **160 (23.6)** | **81 (23.5)** | **0.362** |
|  | **1-2** | **65 (6.9)** | **263 (7.1)** |  | **64 (9.4)** | **21 (6.1)** |  |
|  | **3-4** | **117 (12.4)** | **538 (14.6)** |  | **89 (13.1)** | **49 (14.2)** |  |
|  | **7-8** | **323 (34.2)** | **1165 (31.5)** |  | **231 (34.0)** | **114 (33.1)** |  |
|  | **9-10** | **209 (22.1)** | **781 (21.1)** |  | **135 (19.9)** | **79 (23.0)** |  |
| **Anxiety related to return to lockdown** | **5-6** | **260 (27.5)** | **1063 (28.8)** | **0.346** | **175 (25.8)** | **113 (32.8)** | **0.009** |
|  | **1-2** | **172 (18.2)** | **588 (15.9)** |  | **126 (18.6)** | **40 (11.6)** |  |
|  | **3-4** | **149 (15.8)** | **636 (17.2)** |  | **120 (17.7)** | **49 (14.2)** |  |
|  | **7-8** | **255 (27.0)** | **1018 (27.5)** |  | **180 (26.5)** | **103 (29.9)** |  |
|  | **9-10** | **109 (11.5)** | **391 (10.6)** |  | **78 (11.5)** | **39 (11.3)** |  |
| **Acceptance of Covid-19 vaccine if available** | **Not sure** | **259 (27.4)** | **482 (13.0)** | **<0.001** | **157 (23.1)** | **50 (14.5)** | **<0.001** |
|  | **No** | **152 (16.1)** | **163 (4.4)** |  | **63 (9.3)** | **13 (3.8)** |  |
|  | **Yes** | **534 (56.5)** | **3051 (82.5)** |  | **459 (67.6)** | **281 (81.7)** |  |

Differences in categorical variables assessed using Chi-squared tests.Differences in continuous variables assessed using t-tests.
